# Benign-Malignant Classification of Pulmonary Nodules in CT Images Based on Fractal Spectrum Analysis

**DOI:** 10.1101/2025.08.24.25334331

**Authors:** Yingying Ma, Shichang Lei, Baixin Wang, Yangzhizi Qiao, Fangliang Xing, Tengxiao Liang

## Abstract

This study reveals that pulmonary nodules exhibit distinct multifractal characteristics, with malignant nodules demonstrating significantly higher fractal dimensions at larger scales. Based on this fundamental finding, an automatic benign-malignant classification method for pulmonary nodules in CT images was developed using fractal spectrum analysis. By computing continuous three-dimensional fractal dimensions on 121 nodule samples from the LIDC-IDRI database, a 201-dimensional fractal feature spectrum was extracted, and a simplified multilayer perceptron neural network (with only 6×6 minimal neural network nodes in the intermediate layers) was constructed for pulmonary nodule classification. Experimental results demonstrate that this method achieved 96.69% accuracy in distinguishing benign from malignant pulmonary nodules. The discovery of scale-dependent multifractal properties enables fractal spectrum analysis to effectively capture the complexity differences in multi-scale structures of malignant nodules, providing an efficient and interpretable AI-aided diagnostic method for early lung cancer diagnosis.

## 1. Introduction

### 1.1 Research Background

Lung cancer is one of the malignant tumors with the highest incidence and mortality rates worldwide [1]. Early detection and accurate diagnosis of benign and malignant pulmonary nodules are crucial for improving patient survival rates [2]. With the widespread application of low-dose spiral CT screening, a large number of pulmonary nodules have been detected, but determining the benign or malignant nature of nodules based on CT data remains a challenging task [3]. How to accurately distinguish between benign and malignant nodules has become an urgent clinical problem.

### 1.2 Related Work

Traditional imaging diagnosis mainly relies on subjective judgment by radiologists, leading to inter-observer variability [4]. Although pathological examination provides more accurate judgment, it also presents problems such as invasive sampling, high risk, and inaccurate sampling [5]. In recent years, computer-aided diagnostic systems based on deep learning have made significant progress [6,7], but their “black box” characteristics limit clinical acceptance [8].

Fractal geometry, as a mathematical tool for describing complex structures, has shown unique advantages in medical image analysis [9,10]. Early studies demonstrated that fractal analysis could effectively characterize tissue heterogeneity and structural complexity in various medical imaging applications [11]. Specifically for pulmonary nodule analysis, fractal dimension has been investigated as a quantitative measure of nodule contour irregularities and internal complexity [12,13]. Lin et al. [14] pioneered the application of fractal features based on fractional Brownian motion model for automatic classification of solitary pulmonary nodules, achieving 83.11% accuracy using support vector machine classifiers. Al-Kadi [15] further demonstrated that higher fractal dimensions in CT images of non-small cell lung cancer are associated with advanced stage and greater FDG uptake on PET imaging.

However, current research on fractal feature-based determination of pulmonary nodule benign-malignant classification remains limited to static fractal feature analysis of two-dimensional images, with constrained accuracy [16,17]. Recent advances have begun exploring texture features such as fractal and gray-level co-occurrence matrix features combined with deep learning architectures [18], but comprehensive three-dimensional fractal spectrum analysis has not been systematically investigated. This has led to a temporary stagnation in current research on fractal feature-based discrimination of pulmonary nodule benign-malignant classification.

### 1.3 Research Innovation

This study is the first to apply three-dimensional fractal spectrum analysis to pulmonary nodule benign-malignant classification, extracting multi-scale fractal features through box-counting method and constructing a highly interpretable classification model. Recent methodological advances in multifractal spectrum analysis have demonstrated the potential for precise quantitative evaluation of pulmonary nodule characteristics [19]. Compared to traditional methods, this approach has rich feature dimensions, high computational efficiency, clear physical meaning, and improved accuracy to a clinically practical level.

## 2. Materials and Methods

### 2.1 Dataset

Determining the benign or malignant nature of pulmonary nodules based on CT data requires diagnostic labels, and the LIDC-IDRI dataset currently represents the highest quality open-source dataset available for this purpose [20]. The dataset was derived from the LIDC-IDRI public database (https://www.cancerimagingarchive.net/collection/lidc-idri/), which contains a total of 1012 case scans. However, only 121 cases have definitive benign-malignant annotations available for analysis. The sample size consists of 121 pulmonary nodules, comprising 54 benign and 67 malignant cases, as documented in the diagnostic data file (https://www.cancerimagingarchive.net/wp-content/uploads/tcia-diagnosis-data-2012-04-20.xls). Training was conducted based on the benign-malignant annotation labels from the “Diagnosis at the Nodule Level” field in the tcia-diagnosis-data-2012-04-20.xls file. The CT scan parameters included slice thickness of 1.0mm levels and pixel spacing ranging from 0.6 to 0.8mm, providing high-resolution imaging data suitable for detailed fractal analysis.

### 2.2 Preprocessing Pipeline

The preprocessing pipeline involved systematic extraction and visualization of pulmonary nodule regions of interest (ROIs). ROI information for all nodules was obtained from the XML annotation file (https://www.cancerimagingarchive.net/wp-content/uploads/LIDC-XML-only.zip), which contains detailed spatial coordinates and geometric parameters for each identified nodule. DICOM sub-images of individual nodules were subsequently extracted based on the recorded nodule center coordinates and diameter information, ensuring precise localization and boundary definition. Pulmonary nodule visualization was performed using the MATLAB computing platform to facilitate three-dimensional reconstruction and analysis. A typical example of the three-dimensional visualization process for local pulmonary nodule images is illustrated in Figure 1, demonstrating both the original CT slice and the corresponding 3D mesh representation used for subsequent fractal analysis.

**Figure 1:**
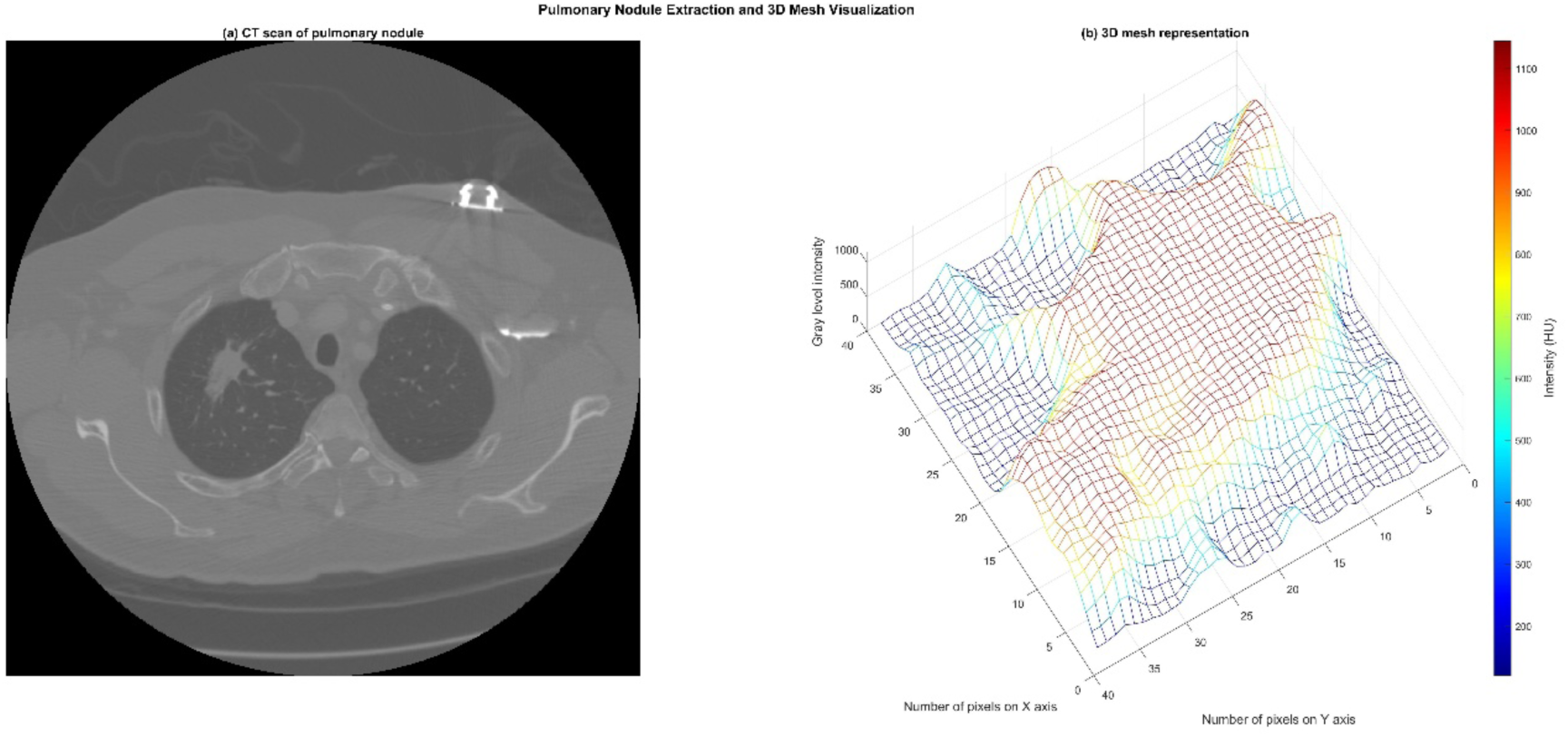
Pulmonary Nodule and Its Gray-Level 3D Visualization.

**Figure 2:**
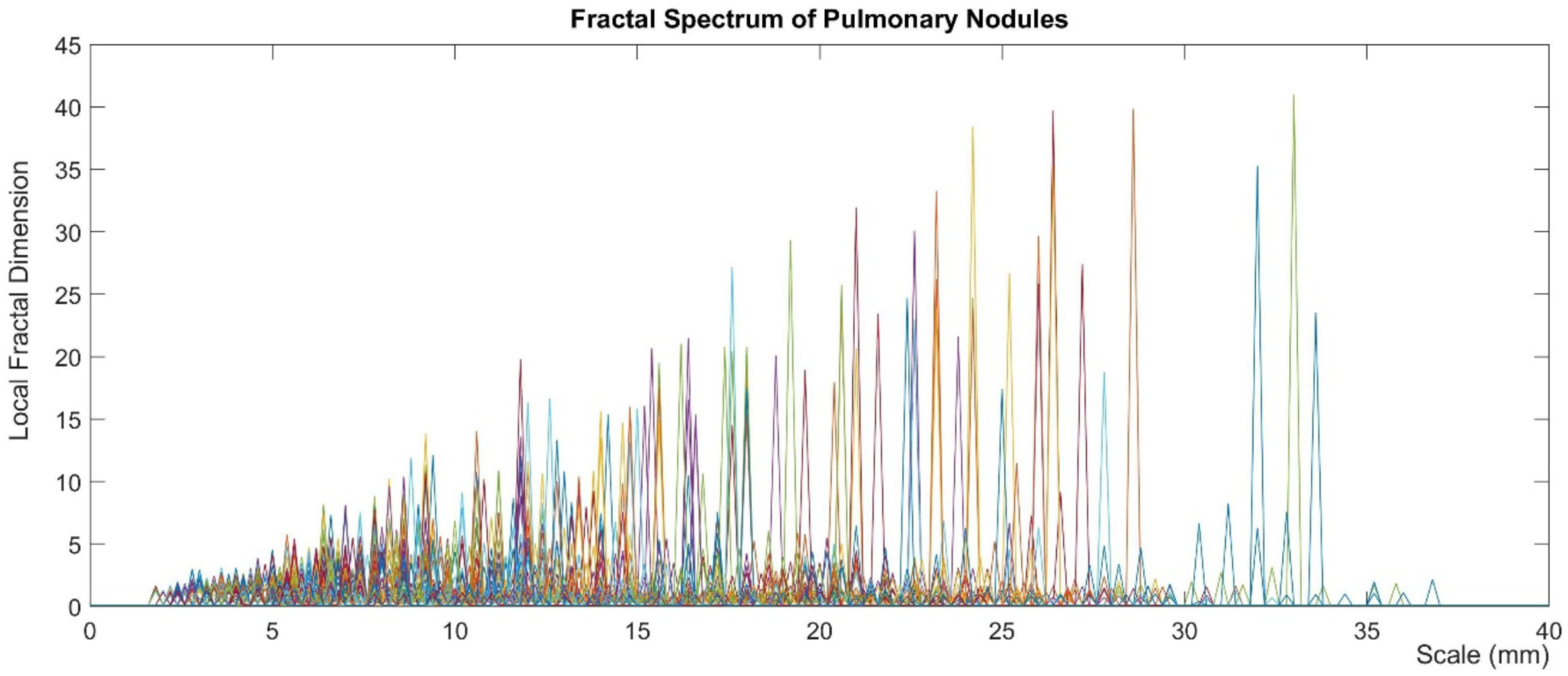
Comprehensive Visualization of Fractal Spectra Across All Pulmonary Nodules.

**Figure 3.**
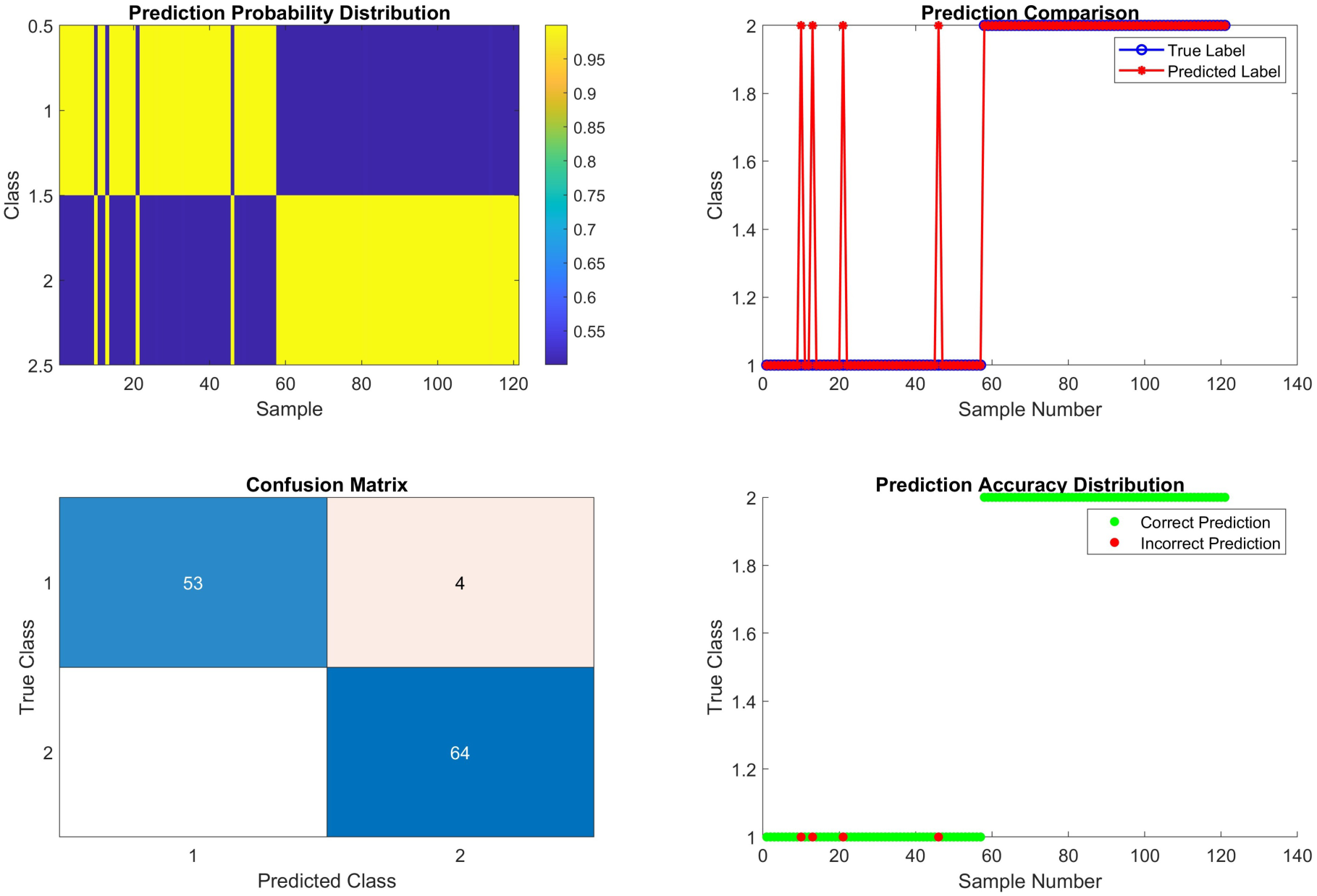
Performance Evaluation of Shallow Multilayer Perceptron for Pulmonary Nodule Classification.

Based on the scanning position of the pulmonary nodule, the corresponding image can be quickly located (Figure 1-a). According to the ROI information, the nodule’s boundaries and corresponding intensity values are extracted to form three-dimensional data (Figure 1-b), where XY represents the two-dimensional pixel directions and Z represents the intensity. Based on this pulmonary nodule 3D mesh, continuous fractal spectrum is calculated.

### 2.3 Fractal Feature Extraction

Fractal dimension is an important parameter describing the complexity of geometric objects [21]. For three-dimensional medical images, fractal dimension D is defined as:

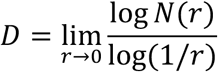

where N(r) is the number of boxes of size r needed to cover the object. The three-dimensional box-counting algorithm calculates local fractal dimensions by counting the number of non-empty boxes N(r) at different scales r:

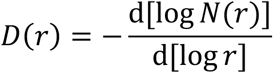

The fractal spectrum reflects the complexity changes of structures at different scales, providing rich feature information for multi-scale structural analysis [22]. During computation, multi-scale analysis was performed in the 0-40mm scale range to calculate local fractal dimensions, using 0.2mm resolution to obtain 201-dimensional unified fractal spectrum features.

### 2.4 Classifier Design

This study employs a shallow multilayer perceptron neural network for pulmonary nodule benign-malignant classification. Choosing the fewest possible neural nodes means that accurate classification based on fractal spectrum features can be achieved without requiring strong nonlinear classification performance, while also ensuring lightweight deployment and efficient computation for clinical practice [23]. The network architecture consists of an input layer (201 nodes), two hidden layers (6 nodes each), and an output layer (2 nodes), forming a four-layer 201-6-6-2 multilayer perceptron structure. Hidden layers use hyperbolic tangent (tanh) activation functions to enhance nonlinear expression capability, while the output layer uses softmax function to ensure probability output. Training employs the Levenberg-Marquardt optimization algorithm with fast convergence characteristics. Key parameter settings include learning rate 0.001, convergence target 1e-8, maximum epochs 50000, maximum failure count 100, and minimum gradient 1e-10, ensuring adequate network training while avoiding overfitting.

**Network Structure:** 201-6-6-2 multilayer perceptron

**Activation Functions:** Hidden layers use tanh, output layer uses softmax

**Training Algorithm:** Levenberg-Marquardt algorithm

**Parameter Settings:** Learning rate 0.001, convergence target 1e-8, maximum epochs 50000, maximum failures 100, minimum gradient 1e-10

Due to the limited sample size (121 cases) and scarcity of annotated pulmonary nodule datasets, all samples were used for training to ensure adequate model learning. The effectiveness of fractal spectrum features was validated through Euclidean distance-based linear classification, providing intuitive verification of feature discriminability based on physical interpretability (Figure 4).

**Figure 4:**
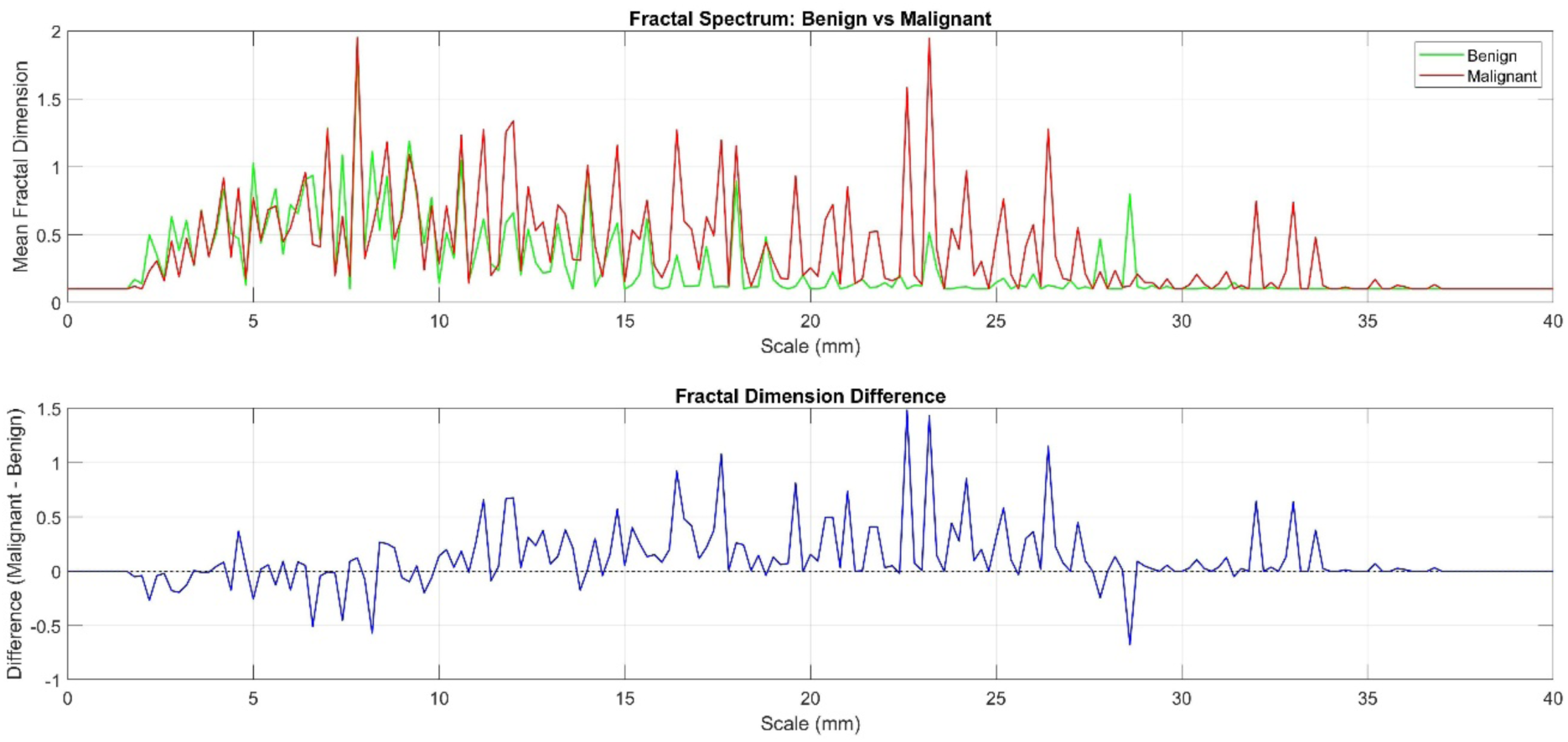
Feature Mean Value Comparison Between Benign and Malignant Nodules.

## 3. Experimental Results

### 3.1 Neural Network Classification Results

The minimalist neural network based on 201-dimensional fractal spectrum features performed excellently in pulmonary nodule benign-malignant classification tasks. The network adopted a 201-6-6-2 architecture, trained through the Levenberg-Marquardt algorithm, achieving high-precision classification on 121 samples.

The shallow multilayer perceptron achieved exceptional classification performance with an overall accuracy of 96.69% (117/121 samples). The detailed performance metrics revealed 92.98% sensitivity for benign nodule recognition (53 out of 57 correctly identified) and perfect 100% specificity for malignant nodule detection (all 67 malignant cases correctly classified). Only 4 samples were misclassified in the entire dataset. The confusion matrix analysis demonstrated that all classification errors involved benign nodules being incorrectly labeled as malignant, while no malignant nodules were misclassified as benign. This asymmetric error pattern indicates that the model exhibits a conservative classification tendency, preferring to err on the side of caution by potentially over-diagnosing malignancy rather than missing true malignant cases, which represents a clinically favorable bias for early cancer detection and patient safety.

The confusion matrix shows that the model has stronger recognition capability for malignant nodules, with 4 benign nodules misclassified as malignant, requiring further clinical validation.

### 3.2 Intuitive Interpretation of Fractal Spectra for Benign and Malignant Pulmonary Nodules

#### 3.2.1 Differences in Fractal Spectra Between Benign and Malignant Nodules

The fractal spectrum of benign nodules is mainly distributed in the low-scale segment (4-10), with lower peaks and concentrated distribution. The fractal spectrum of malignant nodules gradually extends to high-scale intervals (scales 11-26), with higher peaks and broader distribution, reflecting the more complex multi-scale structural characteristics of malignant nodules.

#### 3.2.2 Classification Performance Based on Euclidean Distance

Simply using the Euclidean distance to the green line (benign) and red line (malignant) in Figure 4 to determine nodule nature can also achieve 72.73% recognition accuracy. This means that even without considering the correlation and nonlinearity of fractal spectrum features, simple linear distance can achieve relatively ideal recognition performance. This also indirectly confirms that fractal spectra have significant differences in distinguishing benign from malignant pulmonary nodules.

#### 3.2.3 Comparative Analysis of Fractal Spectra in Typical Benign and Malignant Nodules

Figure 5 demonstrates the comprehensive analysis pipeline for pulmonary nodule characterization, comparing typical benign and malignant cases through multi-dimensional visualization approaches. (a) CT scan of a benign pulmonary nodule showing regular morphology with well-defined boundaries and homogeneous internal density distribution. (b) 3D mesh representation of the benign nodule reveals a smooth, relatively simple surface topology with minimal structural complexity and uniform geometric characteristics. (c) Fractal spectrum of the benign nodule exhibits concentrated distribution primarily within the 5-15mm scale range, with moderate peak values around 10-12 and limited extension to higher scales. (d) CT scan of a malignant pulmonary nodule displays irregular morphology characterized by indistinct boundaries, heterogeneous internal structure, and complex architectural features. (e) 3D mesh representation of the malignant nodule demonstrates significantly increased surface complexity, irregular topology, and pronounced structural heterogeneity with substantial geometric variations. (f) Fractal spectrum of the malignant nodule shows extended distribution across the entire 0-40mm scale range, with significantly higher peak values reaching 40+ and broader spectral coverage, reflecting the enhanced structural complexity and irregular geometric properties characteristic of malignant lesions.

**Figure 5.**
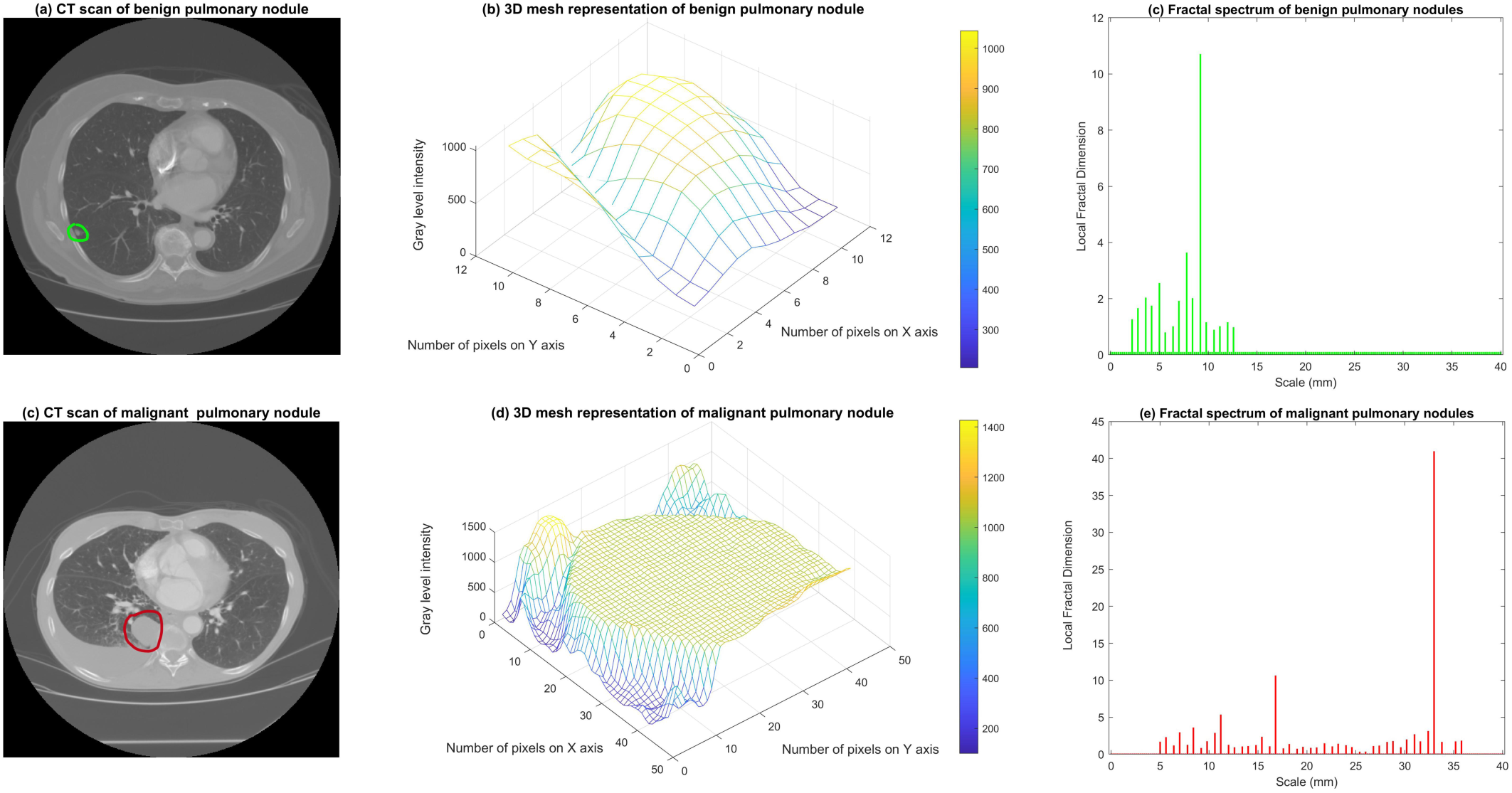
Comparative Analysis of Benign and Malignant Pulmonary Nodules: Multi-Scale Visualization and Fractal Spectrum Characterization.

Figure 5 indicates that pulmonary nodules inherently possess multifractal characteristics, with more malignant nodules exhibiting higher fractal dimensions at larger scales. Perhaps this multifractal property is also present in other types of tumors [24].

### 3.3 LSTM validation

To further validate fractal spectrum discriminative power, an LSTM neural network was implemented using temporal sequence modeling. Unlike the shallow multilayer perceptron treating the 201-dimensional fractal spectrum as static features, the LSTM approach models each spectrum as sequential data, capturing temporal dependencies and transition patterns across scales.

The LSTM architecture consisted of 80 hidden units with approximately 26,000 trainable parameters. Each nodule’s fractal spectrum was reformatted as a time series where scale measurements represented temporal steps, enabling the network to learn sequential relationships between consecutive fractal dimension values. Training employed 30,000 epochs using the Levenberg-Marquardt optimization algorithm with cross-entropy loss function.

The LSTM achieved 96.69% classification accuracy, matching the shallow multilayer perceptron performance. The confusion matrix revealed 55 correctly classified benign cases out of 57 (96.49% sensitivity) and 62 correctly classified malignant cases out of 64 (96.88% specificity). Only 4 samples were misclassified, demonstrating exceptional temporal modeling capability.

The confusion matrix based on the LSTM model is as follows:

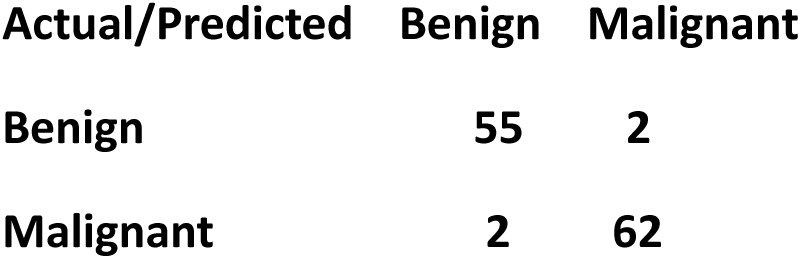

The convergence of results between static (96.69%) and sequential (96.69%) modeling approaches provides strong validation of fractal spectrum methodology. Two fundamentally different architectures achieved identical performance, demonstrating that discriminative power resides within the fractal features themselves rather than specific modeling assumptions, further supporting the clinical utility of three-dimensional fractal spectrum analysis for automated pulmonary nodule classification.

## 4. Discussion

### 4.1 Method Advantages

The proposed fractal spectrum analysis methodology demonstrates several significant advantages for pulmonary nodule classification. The achieved 96.69% classification accuracy represents advanced performance levels, competing effectively with state-of-the-art deep learning approaches while maintaining superior interpretability [25,26]. This high accuracy was consistently achieved across different modeling approaches, including linear classifiers, shallow neural networks, and LSTM architectures, demonstrating the robustness of fractal spectrum features (Sequences of fractal dimensions at different scales).

The strong interpretability of fractal features provides clear physical meaning that facilitates clinical understanding and acceptance [27]. Benign nodules exhibit relatively smaller fractal scales with lower fractal dimensions, reflecting their regular, homogeneous internal structure. In contrast, malignant nodules demonstrate larger fractal scales with correspondingly higher fractal dimensions, indicating their complex, heterogeneous architecture and irregular boundaries [28]. This physical interpretation enables radiologists to understand the mathematical basis for automated decisions. Remarkably, even simple Euclidean distance-based linear classification leveraging this physical understanding achieved 72.73% accuracy, which represents the current highest level in this field using linear methods [29]. The incorporation of minimal nonlinear classification capability through a 12-node shallow perceptron further enhanced recognition rates to 96.69%.

**That pulmonary nodules exhibit multifractal properties constitutes the most important discovery of this research. Leveraging this finding, fractal spectrum analysis significantly enhanced identification accuracy.** Furthermore, this discovery has potential applicability to other tumor types, as recent studies have demonstrated the utility of fractal analysis in various malignancies including cutaneous squamous cell carcinoma and hepatocellular carcinoma [24,30].

The low computational complexity represents a crucial advantage for clinical deployment [31]. Whether employing linear classifiers, 12-node shallow perceptrons, or 80-node LSTM networks, all approaches achieved excellent classification performance while maintaining lightweight architectures suitable for rapid clinical deployment. This computational efficiency enables real-time benign-malignant discrimination without requiring extensive hardware resources or prolonged processing times.

The rapid diagnosis capability addresses critical clinical needs for timely decision-making in pulmonary nodule management [32]. The lightweight nature of all implemented classifiers facilitates seamless integration into existing clinical workflows, supporting immediate classification upon image acquisition.

### 4.2 Clinical Significance

Fractal spectrum analysis provides quantitative characterization of pulmonary nodule structural complexity, offering objective diagnostic evidence for radiologists [33]. This methodology proves particularly valuable for analyzing nodules with indistinct boundaries and irregular morphology, where traditional visual assessment may be challenging and subjective interpretation errors are more likely to occur [34]. The quantitative nature of fractal features eliminates inter-observer variability and provides reproducible measurements that can support consistent diagnostic decision-making across different clinical settings [35].

The clinical implementation of AI tools for lung nodule analysis has shown promising results in supporting radiologists’ decision-making processes [36]. Our fractal-based approach complements existing radiomics methodologies by providing additional quantitative measures that can enhance diagnostic confidence, particularly for challenging cases where conventional morphological features may be ambiguous [37].

### 4.3 Limitations and Improvements

The current study acknowledges several limitations that warrant consideration for future research directions. The primary limitation involves the relatively modest dataset size of 121 cases, which, while sufficient for demonstrating proof-of-concept, necessitates larger-scale validation studies to establish broader clinical generalizability [38]. The sample size constraint may limit the statistical power for detecting subtle performance differences and could affect the robustness of conclusions when applied to diverse patient populations.

The single-center data origin represents another significant limitation, as institutional variations in imaging protocols, patient demographics, and diagnostic practices may influence model performance [39]. Multi-center validation studies encompassing diverse clinical settings, scanner manufacturers, and population characteristics are essential for establishing the universal applicability of fractal spectrum analysis across different healthcare environments.

Feature extension opportunities present promising avenues for performance enhancement. The integration of fractal spectrum features with conventional radiological characteristics, structural analysis parameters, and semantic descriptors could potentially improve classification accuracy beyond the current 96.69% threshold [40]. Multi-modal data fusion represents a particularly compelling improvement strategy, incorporating biochemical markers, genetic profiles, and histopathological information when available [41]. Such comprehensive integration could establish more robust predictive models that leverage the complementary strengths of different data modalities, ultimately advancing toward precision medicine approaches for pulmonary nodule characterization and supporting more informed clinical decision-making in complex diagnostic scenarios.

### 4.4 Comparison with Existing Methods

Compared to pathological biopsy sampling, fractal spectrum analysis offers significant advantages in terms of non-invasiveness and clinical convenience. The method requires only standard CT imaging data without additional invasive procedures, reducing patient risk and discomfort while enabling rapid assessment. This non-invasive approach is particularly beneficial for monitoring nodule progression over time, as repeated evaluations can be performed without patient burden or procedural complications.

The technological advancement trajectory suggests promising future developments. Improvements in CT imaging resolution and enhanced annotation accuracy are expected to further optimize classification performance, potentially achieving near-perfect diagnostic accuracy while preserving the current computational efficiency and interpretability benefits. The integration of fractal analysis into routine clinical workflows could transform pulmonary nodule evaluation from subjective visual assessment to objective, quantitative diagnosis, ultimately improving patient outcomes through more accurate and timely malignancy detection.

## 5. Conclusions

This study reveals that pulmonary nodules inherently exhibit multifractal characteristics, with malignant nodules showing significantly higher fractal dimensions at larger scales. Building upon this fundamental discovery, an automated classification method for benign and malignant pulmonary nodules was successfully established based on three-dimensional fractal spectrum analysis, demonstrating good diagnostic performance and clinical utility. The research makes several significant contributions to AI-aided diagnosis in pulmonary medicine. The methodological innovation represents the first application of comprehensive three-dimensional fractal spectrum analysis to pulmonary nodule classification, achieving 96.69% accuracy that competes with state-of-the-art approaches while maintaining superior interpretability. The proposed 201-dimensional fractal spectrum features effectively capture multi-scale structural complexity, providing quantitative characterization of nodule heterogeneity that correlates with malignancy potential.

The architectural simplification achieved through lightweight neural networks demonstrates that complex diagnostic tasks can be accomplished without extensive computational resources. Both 12-node shallow perceptrons and 80-node LSTM networks achieved exceptional performance while maintaining rapid processing capabilities suitable for real-time clinical deployment. This computational efficiency, combined with the clear physical interpretation of fractal features, facilitates seamless integration into existing clinical workflows and enables immediate diagnostic support upon image acquisition.

The clinical significance extends beyond technical performance metrics, offering radiologists objective, reproducible measurements that eliminate inter-observer variability and support consistent diagnostic decision-making. The non-invasive nature of fractal analysis provides significant advantages over pathological biopsy sampling, enabling repeated evaluations without patient burden while maintaining diagnostic accuracy.

Future research directions will focus on large-scale, multi-center validation studies to establish broader clinical generalizability across diverse healthcare environments. Feature fusion strategies integrating fractal spectrum characteristics with conventional radiological descriptors and multi-modal biomarkers present promising opportunities for performance enhancement. The continued advancement of CT imaging resolution and annotation accuracy is expected to further optimize classification performance, potentially achieving near-perfect diagnostic accuracy while preserving computational efficiency and interpretability benefits, ultimately transforming pulmonary nodule evaluation toward precision medicine approaches.

## Author Contributions

Conceptualization, F.X.; methodology, F.X.; software, F.X.; validation, Y.M., S.L., B.W. and Y.Q.; formal analysis, Y.M. and S.L.; investigation, Y.M. and S.L.; resources, T.L.; data curation, Y.M. and S.L.; writing—original draft preparation, Y.M. and S.L.; writing—review and editing, F.X. and T.L.; visualization, Y.M. and S.L.; supervision, F.X. and T.L.; project administration, T.L.; funding acquisition, T.L. Clinical insights and medical expertise were provided by B.W. and Y.Q. All authors have read and agreed to the published version of the manuscript.

## Funding

This research was funded by the Clinical Research and Achievement Transformation Capacity Enhancement Pilot Project (DZMG-MLZY-23008) from the Dongzhimen Hospital of Beijing University of Chinese Medicine and the Fifth National Traditional Chinese Medicine Clinical Excellent Talents Research Program organized by the National Administration of Traditional Chinese Medicine (http://www.natcm.gov.cn/renjiaosi/zhengcewenjian/2021-11-04/23082.html).

## Data Availability Statement

The data supporting the findings of this study are available from the Lung Image Database Consortium and Image Database Resource Initiative (LIDC-IDRI) at https://www.cancerimagingarchive.net/collection/lidc-idri/.

## Acknowledgments

This research utilized the Multifractal Spectrum V1.0 software tool for assessing pulmonary nodules, developed by Beijing Intelligent Entropy Science & Technology Co., Ltd. The intellectual property rights of this software tool belong to the company. The authors acknowledge the use of this software in the fractal analysis conducted in this study.

## Conflicts of Interest

The authors declare no conflicts of interest.

## Abbreviations

The following abbreviations are used in this manuscript:

CT: Computed Tomography
LIDC-IDRI: Lung Image Database Consortium and Image Database Resource Initiative
ROI: Region of Interest
DICOM: Digital Imaging and Communications in Medicine
MLP: Multilayer Perceptron
LSTM: Long Short-Term Memory AI Artificial Intelligence

## Notes

### Competing Interest Statement

The authors have declared no competing interest.

